# Injection Drug use Practices and HIV infection among People Who Inject Drugs in Kigali, Rwanda

**DOI:** 10.1101/2021.08.05.21261564

**Authors:** Jean Olivier Twahirwa Rwema, Vianney Nizeyimana, Neia Prata Menezes, Nneoma E. Okonkwo, Amelia Aibina Mazzei, Sulemani Muhirwa, Athanase Rukundo, Lisa Lucas, Audace Niyigena, Jean Damascene Makuza, Chris Beyrer, Stefan D. Baral, Aflodis Kagaba

## Abstract

**Background:** In Rwanda, epidemiological data characterizing people who inject drugs (PWID) and their burden of HIV are limited. We examined injecting drug use (IDU) history, practices, and HIV infection in a sample of PWID in Kigali.

**Methods:** From October 2019–February 2020, 322 PWID aged ≥18 were enrolled in a cross-sectional study using convenience sampling in Kigali. Participants underwent a structured interview and HIV testing. We used Poisson regression with robust variance estimation to assess IDU practices associated with HIV infection.

**Results:** The median age was 28 years(IQR:24-31) and 81%(248) were male. The median age at first injection was 23 years (IQR:20-27). HIV prevalence was 9.5%(95%CI:8.7-9.3).

In the six months preceding the study, heroin was the primary drug of choice for 99%(303); but cocaine and methamphetamine were also reported by 10%(31) and 4%(12) respectively. Furthermore, 31%(94) and 33%(103) of participants, shared or reused needles in the previous six months, respectively. Up to 43%(133) knew someone who died from a drug-related overdose. PWID reporting sharing needles at least half the time in the previous six months had increased likelihood of HIV-infection, compared to those who did not (aPR: 2.67; 95%CI:1.23–5.78).

**Conclusion:** HIV infection was common in this sample of PWID in Kigali. The high prevalence of needle reuse and sharing practices highlight significant risk for onward transmission and acquisition of HIV and hepatitis B and C. PWID-focused harm reduction services, including needle and syringe programs, safer injection education, naloxone distribution, and substance use disorder treatment programs, are needed in Rwanda.

## Introduction

Sustained improvements in access to HIV prevention and treatment have been made over the last decade in Eastern and Southern Africa (ESA), the region with the greatest burden of HIV globally ^1^. As a result, since 2010, AIDS-related deaths in the region have fallen by 49% and new HIV infections by 38% ^1^. However, not all people affected or infected by HIV have been reached by these services equally. Key populations (KPs) including men who have sex with men (MSM), transgender women (TGW), female sex workers (FSW) and people who inject drugs (PWID) are not adequately reached with HIV services and remain at a high risk of HIV transmission and acquisition. For instance, HIV prevalence among PWID in ESA is estimated to be around 19%, almost double the prevalence among other adults in the region ^1^.

Globally, 15 million people report injection drug use (IDU), with increasing numbers in countries across sub-Saharan Africa (SSA) ^2,3^. Estimates of IDU range from 0% to 3% among the general adult populations in SSA. IDU is associated with an increased risk of acquiring HIV and other sexually transmitted infections (STIs), hepatitis B (HBV), hepatitis C (HCV), and other blood-borne infections, and is associated with increased burden of mental health disorders ^3,4^. The high risk of HIV among PWID can be explained by individual factors like needle sharing during injection, and structural factors including stigma and discrimination, criminalization, high exposure to human rights abuses; and a paucity of prevention and harm reduction programs ^5,6^. These structural level factors limit PWID engagement in prevention and treatment services, resulting in PWID having poorer HIV outcomes compared to other adults ^7^. PWID additionally face significant social and economic challenges with high burden of homelessness or unstable housing and high incarceration rates ^2^. Finally, PWID have overlapping sexual risks with high levels of inconsistent condom use and sex work ^8^.

While the specific needs of PWID have been widely explored and documented in high-income countries, limited data exist for countries across SSA ^2,9^. However, a review of IDU in six African countries found a high prevalence of high-risk behaviors including needle sharing and inconsistent condom use among PWID ^10^. In addition to the paucity of evidence to guide programing, few PWID-centered programs exist ^9^. A systematic review of interventions to prevent and manage HIV and HCV among PWID revealed that while more countries in SSA are investigating IDU (increased from 13 to 36 between 2007 and 2017), few countries are implementing PWID-focused disease prevention and treatment strategies (2 to 7 for needle and syringe programs, 4 to 8 for opioid substitution therapy, and even fewer for other interventions)^11^. Thus, a greater focus should be placed on PWID programs including epidemiological and clinical research to guide implementation across SSA.

In Rwanda, HIV prevalence among reproductive aged adults has stabilized at 3% for the last decade, with the country being one of the few to achieve the UNAIDS 90-90-90 HIV elimination targets by 2020 ^1^. However, similar progress has yet to be seen among key populations. Recent studies have demonstrated a high prevalence of HIV and other STIs and programmatic gaps among MSM, TGW and FSW ^12-14^. Moreover, PWID have been the most overlooked in HIV programming and research in Rwanda. Rwanda’s HIV guidelines have a minimum package of services for both MSM and FSW, but no such program for PWID^15^. This lack of PWID-focused national programming, coupled with the criminalization of drug use or possession, further complicates programming in Rwanda ^1^. Thus, understanding and addressing the needs of PWID in Rwanda is a public health and human rights imperative.

We conducted this study to better inform future research and programming for PWID in Rwanda. The aim of this paper is to estimate the burden of HIV infection and examine IDU related behaviors associated with HIV among PWID in Kigali.

## Methods

### Study context, procedures, and population

This was a cross-sectional mixed methods study implemented by Health Development Initiative (HDI), a local non-governmental organization working with KPs in Rwanda. This study leveraged qualitative and quantitative methods to guide implementation of health programs for PWID in Rwanda. The specific objectives were to collect data on sociodemographic characteristics and IDU practices among PWID, to characterize the PWID population, and to provide an estimate of the HIV prevalence among PWID in Kigali.

Given the lack of a sampling frame and nonexistence of reliable epidemiological or program data to guide probability sampling methods for PWID, study participants were recruited through purposive and convenience sampling in Kigali city from October 2019-February 2020. Initial participants were recruited from clients who use HDI services in Kigali city. Upon recruitment, these participants were trained and encouraged to recruit their peers who injected drugs in the study. Further community outreach was conducted by HDI community liaisons in all the three districts of Kigali i.e., Nyarugenge, Kicukiro and Gasabo. Eligible participants were at least 18 years old, had injected drugs in the six months preceding the study, agreed to an HIV test, and gave written consent to participate in the study. The study was approved by the Rwanda National Ethics Committee N: 027/RNEC/2020

### Data collection

Data collection was conducted in two HDI sites in Kigali city. After signing the informed consent form, participants underwent structured face-to-face interviews conducted by trained data collectors, and biological testing for HIV. The questionnaire comprised of questions related to sociodemographic characteristics and IDU history and practices in the months preceding the study. Participants were also asked about their need for, and access to, substance use disorder treatment programs. No personal identifying information was collected to protect the privacy of participants.

We performed HIV rapid testing for all participants who gave their consent, per the national guidelines. The screening test was Alere HIV Combo-Determine (Alere, Inc, Waltham, MA) and the confirmatory test was HIV1/2 STAT-PAK (Medford, NY, USA) for participants who screened HIV-positive. Participants with a prior documented HIV diagnosis in a health facility were not retested. Participants who were newly found to be living with HIV were referred to a healthcare facility of their choice to initiate antiretroviral treatment (ART) and further medical management. Participants also received information on centers providing substance use disorder treatment in Rwanda. Upon completion of study procedures, participants received 2000 Frw as transport reimbursement.

### Outcome Assessment

The outcome variable was HIV infection. HIV-negative status was defined only as testing HIV negative at time of the study. HIV-positive status was based on the testing conducted in the study or previously medically confirmed HIV-positive status.

### Other variables of interest

#### Sociodemographic characteristics

Sociodemographic characteristics included age, biological sex, education, occupation, and self-reported sexual orientation.

Age was collected as a continuous variable but categorized into three groups for analytical purposes: 18-24, 25-35 and over 35 years. Sex was defined as female or male based on the sex assignment at birth. Sexual orientation was self-reported and categorized as heterosexual, homosexual, and bisexual. Education and occupation were analyzed as categorical variables with three groups each (Table 1).

**Table 1.** Baseline characteristics of PWID in Kigali, Rwanda, N:307.

| **Characteristic** | **N** | **%** | **Not living with HIV**<br>**N (%)** | **Living with HIV**<br>**N (%)** |
| --- | --- | --- | --- | --- |
| **Age in years** |  |  |  |  |
| 18-24 | 83 | 27.2 | 80 (96.4) | 3 (3.6) |
| 25-34 | 188 | 61.6 | 167 (88.8) | 21 (11.2) |
| >35 | 34 | 11.2 | 30 (88.2) | 4 (11.8) |
| **Biological sex** |  |  |  |  |
| Female | 56 | 18.2 | 48 (85.7) | 8 (14.3) |
| Male | 251 | 81.8 | 230 (91.6) | 21 (8.4) |
| **Education** |  |  |  |  |
| Primary education or less | 47 | 15.3 | 40 (85.1) | 7 (14.9) |
| Some secondary education | 79 | 25.7 | 76 (96.2) | 3 (3.8) |
| Completed secondary or above | 181 | 58.9 | 162 (89.5) | 19 (10.5) |
| **Marital status** |  |  |  |  |
| Single | 252 | 82.9 | 232 (92.1) | 20 (7.9) |
| Cohabiting/Married | 33 | 10.8 | 31 (93.9) | 2 (6.1) |
| Divorced/Separated/Widow | 19 | 6.3 | 12 (63.2) | 7 (36.8) |
| **Occupation** |  |  |  |  |
| Unemployed | 169 | 55 | 154 (91.1) | 15 (8.9) |
| Student | 10 | 3.3 | 10 (100) | 0 (0) |
| Part time/Full time employee | 128 | 41.7 | 114 (89.1) | 14 (10.9) |
| **Self-reported sexual orientation** |  |  |  |  |
| Heterosexual | 215 | 70.3 | 198 (92.1) | 17 (7.9) |
| Homosexual | 35 | 11.4 | 31 (88.6) | 4 (11.4) |
| Bisexual | 56 | 18.3 | 48 (85.7) | 8 (14.3) |

#### IDU history and practices, access to treatment and other behaviors

Participants were asked the age at which they first injected drugs, the drug they first injected, source of the drug, and whether shared needles during their first drug injection. Overall injection history was assessed by estimating the duration of IDU for each participant and lifetime needle sharing history. Participants were also asked if they knew anyone who had died from a drug overdose. Participants were asked detailed questions about their primary drug of injection, other drugs used, and frequency of drug injection in the six months preceding the study. To assess injection frequency, participants were asked the number of times they injected per day, week, month, or year. Frequency of needle sharing, sharing of other injecting materials, and needle reuse were assessed using 5 item scales ranging from never, rarely, half the time, most of the time, and always. We also captured information on injection partnerships (i.e., sex partners, friends, relatives, drug dealer, strangers or other) and history of exchanging sex for drugs (Table 2). Participants were asked about their knowledge and use of substance use disorder treatment programs in Rwanda (Table 3). Finally, participants were asked questions assessing their HIV knowledge, HIV testing history and condom use during sex.

**Table 2.** Injecting drug use history and practices among PWID in Kigali, Rwanda, N:307.

| **Variable** | **N** | **%** |
| --- | --- | --- |
| ***First drug injection*** |  |  |
| **Age at first injection** |  |  |
| Less than 18 | 53 | 17.3 |
| 18-24 | 132 | 43 |
| >25 | 122 | 39.7 |
| **Drug injected the first time** |  |  |
| Heroin | 304 | 99.1 |
| Cocaine | 2 | 0.6 |
| Methamphetamine | 1 | 0.3 |
| **Person who performed the injection the first time** |  |  |
| Self | 134 | 43.7 |
| Friend | 146 | 47.5 |
| Sex partner | 20 | 6.5 |
| Other | 7 | 2.3 |
| **Needle sharing the first time** |  |  |
| No | 234 | 76.2 |
| Yes | 73 | 23.8 |
| **Source of drug the first time** |  |  |
| Bought them from someone | 259 | 84.4 |
| Received them for free | 41 | 13.4 |
| Traded them for sex | 5 | 1.6 |
| Refused to answer | 2 | 0.6 |
| ***Injecting drug history*** |  |  |
| **Duration of drug injection** |  |  |
| Less than 3 | 129 | 42.3 |
| 4-5 years | 105 | 34.4 |
| Over 5 | 71 | 23.3 |
| **Needle sharing history** |  |  |
| Never shared needles | 27 | 8.8 |
| Ever shared needles | 280 | 91.2 |
| ***Injection drug practices in the previous six months*** |  |  |
| **Primary drug of injection in the previous six months** |  |  |
| Heroin | 303 | 98.7 |
| Cocaine | 2 | 0.65 |
| Methamphetamine | 2 | 0.65 |
| **Heroin injection in the previous six months** |  |  |
| No | 5 | 1.6 |
| Yes | 302 | 98.4 |
| **Cocaine injection in the previous six months** |  |  |
| No | 271 | 89.7 |
| Yes | 31 | 10.3 |
| **Methamphetamine injection in the previous six months** |  |  |
| No | 282 | 95.9 |
| Yes | 12 | 4.1 |
| **Drug injected the first time** |  |  |
| Heroin | 304 | 99.1 |
| Cocaine | 2 | 0.6 |
| Methamphetamine | 1 | 0.3 |
| **Used a Drug combination** |  |  |
| No | 212 | 69.5 |
| Yes | 93 | 30.5 |
| **Needle sharing in the previous six months** |  |  |
| Never | 213 | 69.4 |
| Rarely | 60 | 19.5 |
| Half of the time or more | 34 | 11.1 |
| **Needle reuse in the previous six months** |  |  |
| Never | 204 | 66.5 |
| Rarely | 75 | 24.4 |
| Half of the time or more | 28 | 9.1 |
| **Selling sex for Drugs in the previous six months** |  |  |
| No | 204 | 68.5 |
| Yes | 94 | 31.5 |

**Table 3.** Need of and Access to Drug Addiction Treatment programs among PWID in Kigali, Rwanda, N:307.

| **Variable** | **N** | **%** |
| --- | --- | --- |
| **Tried to reduce or quit drug consumption in the previous 6 months** |  |  |
| No | 122 | 39.9 |
| Yes | 184 | 60.1 |
| **Aware of any Drug addiction treatment programs** |  |  |
| No | 205 | 66.7 |
| Yes | 45 | 14.7 |
| Do not know | 57 | 18.6 |
| **Drug Addiction treatment in the previous six months \*** |  |  |
| No | 290 | 95.4 |
| Yes | 14 | 4.6 |
| **Unable to access Drug Addiction treatment in the previous six months** |  |  |
| No | 259 | 84.4 |
| Yes | 48 | 15.6 |
| **Ever enrolled in a Drug Addiction treatment program** |  |  |
| Never been treated | 263 | 85.7 |
| Ever been treated | 21 | 6.8 |
| Refused to answer | 23 | 7.5 |
| **Know someone who died from a drug overdose** |  |  |
| No | 73 | 24 |
| Yes | 133 | 43.8 |
| Don't know | 98 | 32.2 |

## Analysis

We calculated crude estimates, including means and proportions, for sociodemographic characteristics, the outcome of interest and other covariates. Pearson’s Chi-squared tests (χ2) were used to compare demographic and IDU behaviors by biological sex, and an alpha level of 0.05 was used to attribute statistical significance. We applied Lowess smoothed non-parametric regressions to choose appropriate scales for age and duration of injection drug use. Since both variables showed an approximately linear association with HIV infection, they were both analyzed as continuous variables. All other variables were analyzed as binary or categorical variables. Table 4 summarizes all variables hypothesized to be associated with HIV infection and included in our analyses.

**Table 4:** Factors associated with HIV infection among PWID in Kigali, Rwanda, N:307.

|  | **N**<br>**Living**<br>**with**<br>**HIV** | **%**<br>**Living**<br>**with**<br>**HIV** | **Unadjusted PR**<br>**(95% CI)** | **p-**<br>**value** | **Adjusted PR**<br>**(95%CI)** | **p-**<br>**value** |
| --- | --- | --- | --- | --- | --- | --- |
| **Sociodemographic characteristics** |  |  |  |  |  |  |
| Age in years |  |  | **1.05 (1.01-1.09)** | **0.02** | 0.98 (0.91-1.07) | 0.769 |
| **Biological sex** |  |  |  |  |  |  |
| Female | 8 | 14.3 | Ref |  |  |  |
| Male | 21 | 8.4 | 0.58 (0.27-1.25) | 0.169 |  |  |
| **Education** |  |  |  |  |  |  |
| Primary education or less | 7 | 14.9 | Ref |  | Ref |  |
| Some secondary education | 3 | 3.8 | **0.25 (0.07-0.94)** | **0.04** | 0.31 (0.09-1.05) | 0.06 |
| Completed secondary or above | 19 | 10.5 | 0.71 (0.31-1.57) | 0.395 | 0.64 (0.27-1.50) | 0.303 |
| **Marital status** |  |  |  |  |  |  |
| Single | 20 | 7.9 | Ref |  | Ref |  |
| Cohabiting/Married | 2 | 6.1 | 0.76 (0.18 - 3.13) | 0.708 | 0.81 (0.21-3.26) | 0.769 |
| Divorced/Separated/Widow | 7 | 36.8 | **4.64 (2.25 - 9.58)** | **0.001** | **4.56 (1.91-10.90)** | **0.001** |
| **Occupation \*** |  |  |  |  |  |  |
| Unemployed | 15 | 8.9 | Ref |  |  |  |
| Employed | 14 | 10.9 | 1.23 (0.62-2.46) | 0.554 |  |  |
| **Self-reported sexual orientation** |  |  |  |  |  |  |
| Heterosexual | 17 | 7.9 | Ref |  |  |  |
| Homosexual | 4 | 11.4 | 1.44 (0.52-4.05) | 0.484 |  |  |
| Bisexual | 8 | 14.3 | 1.81 (0.82-3.97) | 0.141 |  |  |
| **Drug Injection related behaviors,**<br>**access to needles and condom use** |  |  |  |  |  |  |
| **Age at first injection** |  |  |  |  |  |  |
| Before 18 years | 5 | 9.4 | Ref |  |  |  |
| 18 to 24 years | 12 | 9.1 | 1.36 (0.57-3.23) | 0.48 |  |  |
| Over 25 years | 12 | 9.8 | 1.82 (0.75-4.37) | 0.183 |  |  |
| **Duration of drug injection** |  |  |  |  |  |  |
| Years injecting drugs |  |  | **1.06 (0.99-1.11)** | **0.055** | 1.05 (0.93-1.18) | 0.407 |
| **Needle sharing in the previous 6 months** |  |  |  |  |  |  |
| Never | 15 | 7.1 | Ref |  | Ref |  |
| Rarely | 6 | 10 | 1.42 (0.57-3.51) | 0.447 | 1.39 (0.48-4.04) | 0.536 |
| Half the time or more | 8 | 23.5 | **3.34 (1.53-7.28)** | **0.002** | **2.67 (1.23-5.78)** | **0.013** |
| **Number of needle-sharing partners in the previous 6 months** |  |  |  |  |  |  |
| None | 7 | 11.1 | Ref |  |  |  |
| One to two | 15 | 8.2 | 0.74 (0.31-1.73) | 0.484 |  |  |

Bivariable Poisson regression models with robust variance estimation were fitted to compute prevalence ratios (PR) and 95% confidence intervals (CI) determining the association between HIV infection and sociodemographic factors and IDU practices. Poisson regressions were used after log binomial models failed to converge. The final model was constructed using variables that were associated with the outcome if P<0.1 in the bivariable analyses. All the analyses were performed with Stata Version 14.2 (StataCorp, College Station, TX) statistical package.

## Results

### Sociodemographic characteristics and HIV infection

A total of 322 PWID were recruited in the study, but these analyses were restricted to 307 participants for whom HIV status information was available (five participants refused testing and 10 were missing testing results). The median age of participants was 28 years (IQR:24–31) and 81% (248) were male. All sociodemographic characteristics are summarized in table 1.

The prevalence of HIV in this group was 9.5% (95%CI: 8.7-9.3).

### Injection drug history

Median age at first injection was 23 (IQR:20-27), but 17% (53) of participants had their first injection before reaching 18. The majority of participants, 57% (176) had been injecting for four or more years. Nearly all, 99% (304), injected heroin their first time and 24% (73) shared needles with someone at their first injection.

In the six months preceding the study, heroin was the primary drug of choice for 99% (303) of participants. However, 10% (31) and 4% (12) of participants, respectively, reported injecting cocaine and methamphetamine as well. Many participants, 30% (93), had used a drug combination in the six months before the study. The most common combination of drugs was heroin and marijuana, reported by 23% (70) of participants, while alcohol use was reported in combination with heroin by 9% (28) of participants. IDU frequency was high with 95% (293) reporting at least one injection per day in the previous six months.

Overall, 31% (94) of participants shared needles while 27% (83) shared other injection paraphernalia in the previous six months. However, up to 91% (280) of participants reported ever sharing needles in their lifetimes. Furthermore, 33% (103) reported reusing needles previously used for injecting. Most participants, 98% (301) reported getting sterile needles from pharmacies and 20% (61) reported also getting needles from their networks, including friends and sex partners, among others. However, 34% (101) of participants reported not always having access to sterile needles for injection purposes when they needed them. Finally, 31% (94) of participants reported exchanging sex for drugs in the six months before the study (Table 2). Females were more likely than men to report exchanging sex for drugs (60% vs 25%, χ^2^ p value < 0.0001). There were no other significant differences between males and females on other variables assessing IDU practices and history.

The majority of participants, 60% (184), reported having tried to reduce or quit IDU in the previous six months. However, only 5% (14) had enrolled in a treatment program within this timeframe. Overall, 43% (133) of participants knew someone who died from a drug-related overdose (Table 3).

Regarding HIV related knowledge and behavior, 87% (268) reported knowing that IDU was a risk factor for HIV infection. Over half of participants, 65% (198), reported inconsistent condom use in the six months preceding the study.

### Bivariable analysis

In the bivariable analyses, sociodemographic characteristics among PWID that were significantly associated with HIV were age, education, and marital status. Each additional year increase in age was associated with a 5% increase in HIV infection (PR: 1.05; 95% CI: 1.01–1.09). Additionally, PWID with secondary education had lower prevalence of HIV (PR: 0.25; 95% CI: 0.07–0.94) compared to those with primary education or less. PWID who were divorced, separated, or widowed had a four times higher burden of HIV compared to individuals who were single (PR: 4.64; 95% CI: 2.25–9.58). There were no significant associations between HIV infection and biological sex, occupation, or sexual orientation in this group (Table 4).

Duration of drug injection and needle sharing in the six months preceding the study were both significantly positively associated with HIV infection. Each additional year of drug injection was associated with a 6% increase in HIV infection (PR: 1.06; 95% CI: 0.99–1.11). Additionally, PWID who shared needles half the time or more in the six months preceding the study were three times more likely to be living with HIV compared to those who had not shared needles during that time frame (PR:3.34; 95% CI: 1.53–7.28). There were no significant associations between HIV and age at first injection, number of sharing partners, or selling sex in the six months preceding the survey (Table 4).

### Multivariable analysis

In the final multivariable model, sharing needles half the time or more during drug injection remained positively associated with HIV infection (aPR: 2.67; 95% CI: 1.23–5.78). (Table 4)

## Discussion

This study is one of the first to characterize the population of PWID in Rwanda and to describe the burden of HIV in this community. Moreover, it clearly demonstrates existence of individual risk factors known to be associated with HIV, including needle sharing and inconsistent condom use. These practices highlight substantial potential risks of onward transmission and acquisition of HIV and other bloodborne infections among PWID in Rwanda. These results show an urgent need for implementation of evidence-based harm reduction strategies and other individual-, network-, and structural-level interventions to reduce morbidity and mortality among PWID in Rwanda.

In this convenience-based sample, PWID carried a disproportionate burden of HIV over three times what is observed in the general population. ^16^ These findings are consistent with other African studies, underscoring the need to address the HIV prevention and treatment needs of PWID to close HIV elimination gaps ^17^. However, other countries have reported higher HIV burden among PWID (e.g., Kenya: 14-20%; Mozambique: 20-50%; South Africa: 21%), likely reflecting the overall higher burden of HIV in these countries compared to Rwanda ^18-21^. Injection partners with whom one shares injection equipment, is a primary mode of HIV acquisition. However, risk of HIV and other bloodborne infections can be further compounded by sexual risk behaviors (e.g., condomless sex). Our results suggest that consistent condom use in this population is low, which is expected given that research suggests riskier sexual practices are associated with substance abuse ^22^. Globally, studies indicate that sexual risks are elevated among PWID in sub-Saharan Africa relative to other regions, highlighting the importance of understanding the intersecting impact of sexual and injection drug risks among PWID in Rwanda^17^. Implementing sexual health programs, like condom distribution and HIV/STI management, could mitigate the added sexual risks that PWID face. Successes of the Rwanda HIV program in the general adult population could be leveraged to develop initiatives tailored to PWID needs.

Importantly, we observed a high prevalence of risky IDU practices. Nine out of ten participants reported ever sharing needles, while over one third reported sharing needles in the prior 6 months. Consistent with existing literature from the region, needle sharing was positively associated with HIV infection in this study ^18,19,21^. A high proportion of participants also reported reusing needles, which is known to be associated with severe infections including abscesses, septicemia, and infective endocarditis ^23-25^. Taken together, these data demonstrate the urgency for implementing comprehensive harm reduction interventions in Rwanda, particularly needle and syringe service programming (NSP) to provide safe injection equipment and education on safer injection techniques ^26^. NSPs providing sterile needles and other drug paraphernalia have been shown to effectively reduce unsafe injection practices and injection frequency, to facilitate linkage to substance use disorder treatment programs and other Medical Assisted Treatment (MAT), and to be cost effective ^27-29^.

In addition to NSP, there is a necessity for overdose prevention and management and substance use disorders treatment in Rwanda. Despite Naloxone already being available in some pharmacies in Rwanda ^30^, nearly half of participants reported knowing someone who died from a drug-related overdose. This calls for immediate introduction of overdose prevention and treatment programs and strategies to optimize distribution and use of Naloxone in Kigali. Community-based overdose prevention and response programs have been implemented in other settings, and there is evidence that these programs can save lives ^31-33^. Finally, over half of participants reported wanting to reduce or quit drug injection practices, however only 15% reported ever attending a substance use treatment program, highlighting a need for increased coverage and availability of programs such as MAT. Heroin was the primary drug of choice, as observed in other countries in the region, unlike in the United States or other Western countries where synthetic/pharmaceutical opioids, often acquired as prescription medication, are injected with higher frequency ^3^. Additionally, some participants reported injecting cocaine and methamphetamine. It is possible that there are more drug types being used in Kigali that are not reported in this study, most likely because participants were not asked about them during interviews. Given the high frequency of injection reported (i.e., most participants injected daily), and the likely high number of overdoses, a comprehensive study evaluating the types of drugs available in Kigali and their chemical composition is needed to inform programming for PWID in Kigali.

Another contribution of this study is characterizing the demographic characteristics of PWID in Kigali. Almost one fifth of participants reported their first injection before 18 years of age. The 2020 World Drug Report highlighted the growing demand for injection drug use, particularly among young adults in African countries ^3,34^. This is evidenced by the fact that almost half of participants in our study reported injecting drugs for fewer than 3 years. Additionally, although there was no statistical association between biological sex and HIV infection, female PWID in our study reported a higher prevalence of selling sex for drugs. Gender differences in the risk of HIV acquisition among PWID have been documented in other settings ^35,36^. Further studies detailing social and health experiences of different demographic subgroups of PWID in Rwanda are needed.

The high prevalence of selling sex among female PWID in this study is consistent with findings from other studies that have documented high levels of soliciting sex in exchange for drugs among female PWID ^36,37^. Additionally, the fact that some PWID in the sample self-identify as MSM suggest existence of groups of PWID who may have substantial sexual risk of HIV and other STI compared to other PWID in Rwanda. For instance, PWID who also engage in sex work have been found to be at higher risk of HIV compared to other PWID ^8^. Thus, programs should be cognizant of this complex interplay of sexual identities; and sexual and IDU practices to offer optimal interventions to all members of KP groups.

Our study has several limitations. First, our sample is cross-sectional and convenience-based. We are therefore unable to make inferences to the larger PWID population in Kigali. Furthermore, the cross-sectional nature of the study limits our ability to make temporal inferences between the variables of interest and HIV infection. Second, separate from HIV testing, our measurers are self-reported and may be subject to recall or social desirability bias. However, we observed high prevalence of injection and needle sharing behaviors, thus providing a benchmark with which to inform programmatic planning for PWID in Rwanda. Our study was also limited in that it did not collect information on the behaviors of injection partners. The importance of injection drug networks on the transmission and acquisition of bloodborne infections, including HIV, and the social diffusion of behaviors and other network norms, have been well documented in other settings ^38^. Given that over half of participants reported injecting with someone else at first injection, indicates the potential for implementing network-based interventions to reach PWID with services and educational messages early in their injection history. Finally, while the study identifies a proportion of the sample that are aware of their HIV status, we are unable to determine HIV outcomes further downstream of the HIV care continuum. In particular, viral suppression could serve as a marker to gauge the success of the HIV treatment program in this population. The lack of HBV and HCV testing in this study presents another limitation given the high burden of these infections among PWID globally ^17^. Despite these limitations, our study provides critical information to understand the HIV programming needs of PWID in Kigali.

Future research should leverage respondent-driven sampling, or other sampling approaches used to reach and estimate the size of historically hidden populations without a known sampling frame ^39,40^. Stronger sampling approaches could facilitate data collection for larger epidemiological studies, enabling estimation of the prevalence of HIV, HBV, HCV, and other health (e.g., viral suppression) and social outcomes (e.g., mental health), as well as the impact of structural factors (e.g., stigma, criminalization) on these outcomes at the community or population level. Estimating the size of the PWID population would allow for better programmatic planning to address the HIV and other health needs of PWID in Rwanda.

## Conclusion

The findings of this study call for immediate action. PWID have a high HIV prevalence and self-reported injection practices associated with substantial onward risk of transmission and acquisition of HIV and other bloodborne infections. Implementation of evidence-based comprehensive harm reduction programs is not only a public health emergency but also a human rights and moral imperative. Few African countries have adopted needle and syringe exchange programs, and even fewer have government-led MATs. Rwanda should join this list to save lives of PWID and generate evidence that will inform programming in other African countries.

## Data Availability

The data can be made available upon reasonable request. Requests for data utilization should be sent to Dr. Aflodis Kagaba

## Acknowledgments

We are grateful to all study participants for their time and for sharing their personal histories with us.

## Authors’ contribution

VN, AAM, SM, AR, AK, AN conceived the study. SM, VN, AK led the implementation of the study. JOTR, NPM, NO analyzed the data. JOTR wrote the first draft of the paper. All authors edited and approved the manuscript.

